# Economic evaluation of delaying the infant hepatitis B vaccination schedule

**DOI:** 10.1101/2025.11.24.25340907

**Authors:** Eric W. Hall, Prabhu Gounder, Heather Bradley, Noele P. Nelson

## Abstract

**Introduction:** Children who acquire hepatitis B virus (HBV) infection in early childhood through perinatal, household or community exposures are at highest risk of all age groups for experiencing chronic infection and premature death. Universal administration of hepatitis B (HepB) vaccine for all infants <24 hours of birth and completing the childhood series remains the cornerstone for HBV elimination efforts in the United States. We evaluated the health and economic impact of delaying HepB vaccination among U.S. infants.

**Methods:** We constructed a Markov model to simulate lifetime health outcomes and costs for the 3,628,934 infants born in the United States in 2024 for HBV infections acquired up to age 18 years. The model incorporated hepatitis B surface antigen (HBsAg) and hepatitis B e antigen (HBeAg) prevalence among birthing parents, percentage of pregnant parents tested for HBsAg during pregnancy or at delivery, vertical/perinatal and household/community HBV infection risks among children, and resulting sequalae from chronic infection. Eight delayed vaccination scenarios were evaluated in which the first HepB dose was given at 2 months, 7 months, 4 years, or 12 years, applied either only to infants born to HBsAg-negative birth parent or to both HBsAg-negative and HBsAg-unknown birthing parents. Outcomes included acute and chronic HBV infections, cirrhosis, hepatocellular carcinoma, HBV-related deaths, quality-adjusted life-years (QALYs), life-years, and total healthcare costs from the healthcare system perspective. We modeled scenarios with perfect and imperfect adherence to the HepB vaccination schedule, and scenarios with or without additional HBV screening costs for those with delayed receipt of HepB vaccine.

**Results:** All delayed vaccination scenarios resulted in more infections, worse health outcomes, and higher costs than the current universal birth dose recommendation. Under perfect adherence, delaying HepB vaccination by 2 months for infants of HBsAg-negative parents led to an additional 90 acute infections, 75 chronic infections, 29 HBV-related deaths, with $16.4 million in added costs for infants born during one year. Delaying to 12 years resulted in an additional 190 acute infections, 50 deaths, and nearly $30 million in added costs. Delaying HepB vaccination to 12 years for infants of both HBsAg-negative and HBsAg-unknown parents resulted in an additional 2,351 acute infections, 744 deaths, and $368 million in excess costs. Imperfect adherence to the vaccination schedule amplified all negative outcomes substantially. Incorporating pre-vaccination serologic screening for delayed schedules markedly increased total costs.

**Conclusions:** Even brief delays in HepB vaccine initiation substantially increase HBV infections, adverse health outcomes, and health system costs. Our results quantify and demonstrate the importance of the universal HepB birth dose in preventing perinatal and early childhood HBV transmission in the United States.

**Summary:**

- We constructed a Markov model to evaluate health and economic outcomes for the 3.6 million infants born in 2024 under eight scenarios that delayed the first hepatitis B (HepB) vaccine dose to 2 months, 7 months, 4 years, or 12 years.
- All delayed schedules increased acute and chronic infections, HBV-related deaths, and healthcare costs compared with the current universal HepB birth-dose policy.
- For a single year of delaying the HepB birth dose to 2 months among infants whose birth parents are not known to be living with hepatitis B, we estimate 1,437 preventable hepatitis B infections among children, 304 cases of liver cancer, 482 HBV-related deaths, and over $222 million in excess healthcare costs.
- For a single year of delaying the HepB birth dose to 12 years among infants whose birth parents are not known to be living with hepatitis B, we estimate 2,726 preventable hepatitis B infections among children, 503 cases of liver cancer, 788 HBV-related deaths, and over $313 million in excess healthcare costs.
- Our results demonstrate and quantify the critical role of timely universal HepB birth dose vaccination in preventing childhood HBV transmission.

## Introduction

Chronic hepatitis B virus (HBV) infection is a leading cause of cirrhosis, liver failure, liver cancer, and death that affects an estimated 2.4 million persons in the United States.^1,2^ Approximately 70% of persons with chronic HBV were born outside the United States.^3^ The risk of developing chronic infection is age-dependent and highest with perinatal HBV transmission; up to 90% of infants with acute HBV infection will progress to chronic infection compared with 5% of adults. Chronic HBV is incurable, and 25% of children with perinatally acquired infection will die prematurely from cirrhosis or liver cancer.^4-7^ Therefore, preventing HBV transmission at birth remains a cornerstone of HBV elimination efforts.

While perinatal transmission is the most common and severe mode of HBV transmission among infants, horizontal transmission via household or community contact is a mode of transmission that occurs more often with increasing chronic HBV prevalence.^8-10^ HBV survives outside the human body for at least 7 days^11^, and can be transmitted to children via contaminated objects or environmental surfaces at home, in day care, school, or other settings.^12^ Children less than 5 years of age with HBV infection, have about a 30% risk of progression to chronic infection.^13^

Following the licensure of Hepatitis B (HepB) vaccines in the United States in 1982, a series of progressively more comprehensive screening and vaccination recommendations were adopted to prevent perinatal HBV transmission. Initially, prenatal HBV screening was only recommended for pregnant persons with risk factors for infection. Infants born to known hepatitis B surface antigen (HBsAg) birth parents were recommended to receive HepB vaccine and hepatitis B immune globulin (HBIG) at birth, which reduced the risk of perinatal HBV transmission by 94%. Because risk factor based screening failed to identify all HBsAg positive pregnant parents, ACIP recommended prenatal screening of all pregnant women in each pregnancy in 1988.^14^ Gaps in implementation of the screening recommendations resulted in missed opportunities for identifying HBsAg+ pregnant persons, which prompted ACIP to recommend universal infant hepatitis B vaccination in 1991.

Perinatal HBV infections continued to occur for various reasons including the use of incorrect screening tests, errors in interpreting the screening test or the transcription of the screening test, lapses in providing standard of care postexposure prophylaxis, and pregnant persons with false negative test results when conducted during the acute/window period of HBV infection.^15,16^ Therefore, ACIP recommended HepB vaccine for all infants within 24 hours of birth in 2018.^16^

In addition to the recommendations for screening of pregnant women and offering HepB vaccine at birth, ACIP recommended adolescent catch-up vaccination in 1995 and universal vaccination of all children childhood (≤18 years) in 1999 to reduce childhood HBV infections.^17,18^ From 1990 through 2019, cases of acute hepatitis B declined by 99% (from 2,180 to 13) among people aged 0-19 years, demonstrating the remarkable success of the United States vaccination program.^19,20^ Through continued rigorous implementation of HBV screening/testing, HepB vaccination and antiviral treatment, HBV transmission can be eliminated in the United States. The potential for elimination is exemplified by the Alaska Native tribal health system’s vaccine program. Following a statewide HBV screening and vaccination campaign in the early 1980s, the Alaska Native tribes implemented a policy of universal infant and childhood HepB vaccination. By 2008, there were only two known

HBsAg positive children aged <20 years and no transmission of HBV in the Alaska Native population.^12^ The universal HepB vaccine birth dose within 24 hours of birth (followed by a complete vaccine series) is the global standard recommendation for prevention of perinatal and early childhood HBV transmission.^21,22^ This approach is the foundation for HBV elimination efforts and to protect infants and children from the devastating sequalae of HBV infection. Any change to the universal birth dose recommendation poses a serious risk of removing the safety net for infant and childhood prevention of HBV infection and unravelling the strategy to eliminate HBV transmission in the United States. We conducted an economic evaluation of changing the current birth dose recommendation and delaying the initiation of HepB vaccination among infants in the United States.

## Methods

### Cohort

We modeled health and economic outcomes among a cohort of 3,628,934 infants, based on the number of births that occurred in the United States in 2024.^23^ Prior to simulating the cohort of infants, we modeled HBsAg/HBeAg status and prenatal care among the pregnant parents. Utilizing data from a large retrospective analysis of women of childbearing age, we assumed 0.47% of birthing parents were infected with a chronic HBV infection (HBsAg+) at the start of pregnancy^24^, of which 35.8% were HBeAg+.^25^ Based on data that report 14.6% of pregnancies receive either no or inadequate prenatal care (defined as after the 4^th^ month or fewer than 50% of recommended visits)^26^ and that 14.6% of pregnancies which receive adequate prenatal care do not receive HBsAg testing^27^, we assumed 72.9% of pregnancies received prenatal HBsAg testing in the third month of pregnancy. After modeling HBsAg prevalence and probability of receiving prenatal HBsAg testing at month 3, we categorized all pregnant parents into three categories: HBsAg test-positive, HBsAg test-negative, and HBsAg test-unknown. Pregnant persons that were not HBsAg+ at month 3 were assumed to not be immune or receive vaccine during pregnancy and would experience a monthly risk of acute HBV infection of 0.00005, which was calculated using an estimate of acute HBV prevalence among women of childbearing age (0.03%)^24^ and an assumption that IgM anti-HBc remains detectable for 6 months (monthly probability: p=1−(1−0.0003)1/6=0.00005). All acute infections that occurred during pregnancy were assumed to be HBeAg+ at birth. Of the pregnancies that did not receive prenatal HBsAg testing, 20.9% (range: 19.1%-22.8%) were tested at delivery.^28^ We assumed all HBsAg testing had a 99% sensitivity and 100% specificity.^29,30^

### Risk of HBV infection and disease progression

Using a Markov model with a time step of one month, we quantified health and economic outcomes for the lifetime of the cohort of infants. Importantly, this analysis focused on changes to the child and adolescent vaccine schedules and assumed any changes would not impact adult HepB vaccine recommendations. Therefore, we only quantified diffrences in vaccination and health outcomes related to HBV infections that are acquired during childhood and adolescence (i.e. prior to 18 years of age) and we did not model incident HBV infections that may be acquired later as adults. Without vaccination or HBIG at birth, 85% of infants born to HBsAg+/HBeAg+ and 30% born to HBsAg+/HBeAg-pregnant parents acquired an acute HBV infection perinatally.^16^ Susceptible children born to an HBsAg+ parent that were not infected during the perinatal phase faced a risk of acute HBV infection through household transmission.^10^ In our model, these infants have a 45% (range: 30%-60%) chance of becoming infected by 5 years of age.^9,31^ We converted this cumulative probability to monthly risk using the formula r=−(ln(1−P))/t. A previous study of school children in the United Kingdom estimated an annual incidence of acute HBV of 14.99/100,000 among school children 7-11 years old^32^. We converted this to a monthly probability and applied it to children born to an HBsAg+ parent that are still susceptible to infection from age 6 through 11 years. Finally, all susceptible children, regardless of pregnant parent HBsAg status, face a risk of acute HBV infection through community transmission. Given the large-scale success of infant vaccination in the United States, data are limited on the risk of community transmission for children and adolescents. We assumed an annual risk of infection of 0.01% between ages 12 and 18 years.

A proportion of individuals that acquire an acute HBV infection progress to a stage of chronic HBV infection. Chronic HBV infection occurred in 90% of children that were infected during infancy (i.e. <1 year) and 30% of children infected before 6 years^13,16,33^. Similar to previous models of perinatal HBV infection, we assumed all chronic infections acquired before 6 years started in the immune tolerant phase and experienced monthly transition probabilities so that by 20 years of age, 8% of patients with a perinatal infection cleared HBsAg, 45% achieved inactive carrier status, 45% transitioned to active HBV, 1% developed HCC, and 1% developed cirrhosis.^34^ Among children and adolescents that acquire an acute infection between 6 and 18 years of age, 6% (range: 1%-12%) progressed to a chronic infection in the active HBV state.^16^ At 18 years of age, subsequent disease progression rates were from a previously published and validated model of HBV progression among adults^35^. We assumed 49.8% (range: 25.1% - 74.6%) of persons with a chronic HBV infection were diagnosed^2^, of which 36% were linked to care and 18% initiated treatment^36^ at 45 years of age. All health states and annual transition rates are reported in Table 1. All ages and health states experienced mortality defined by annual probabilities of death from 2022 US Census Life Tables.^37^

**Table 1.**
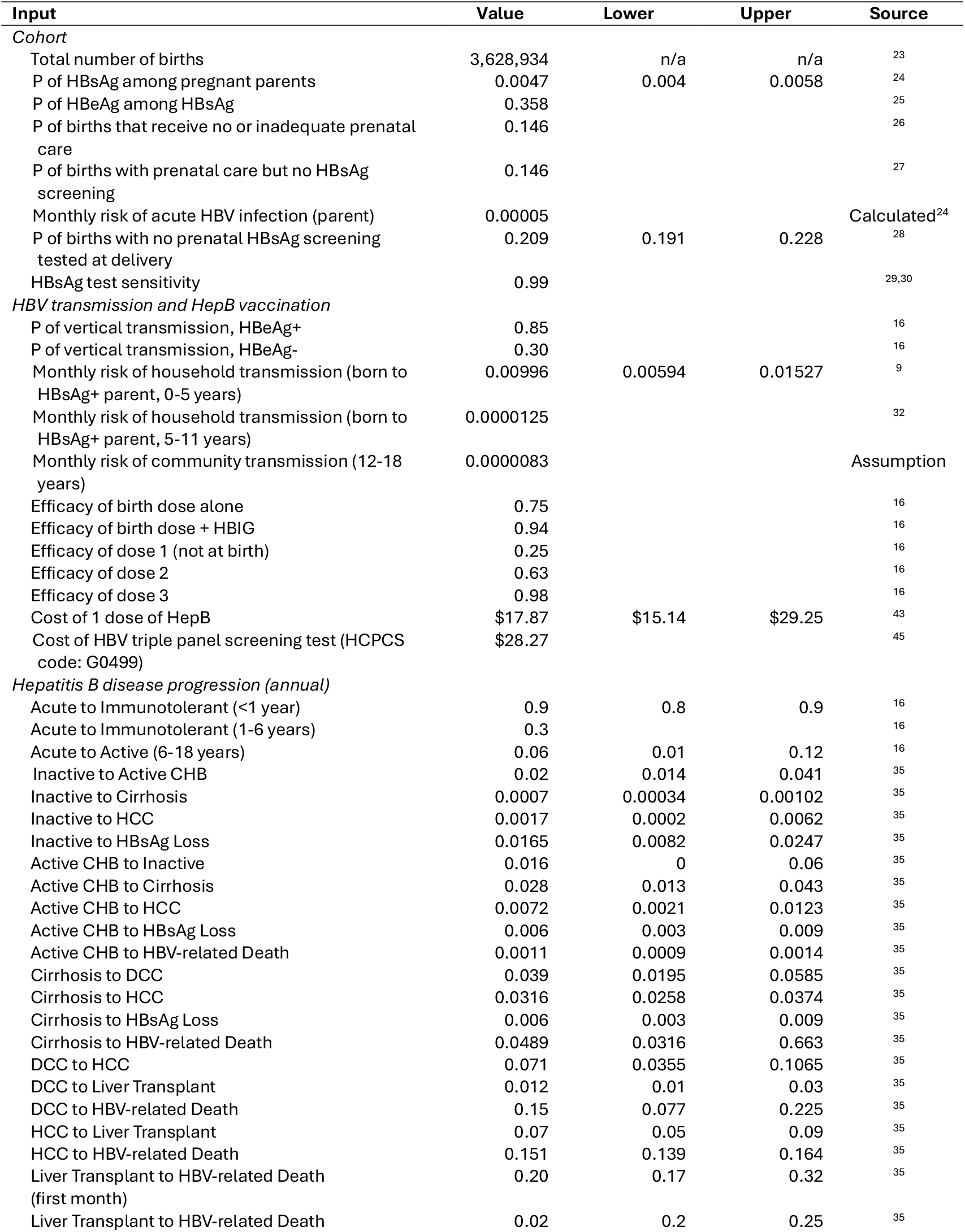
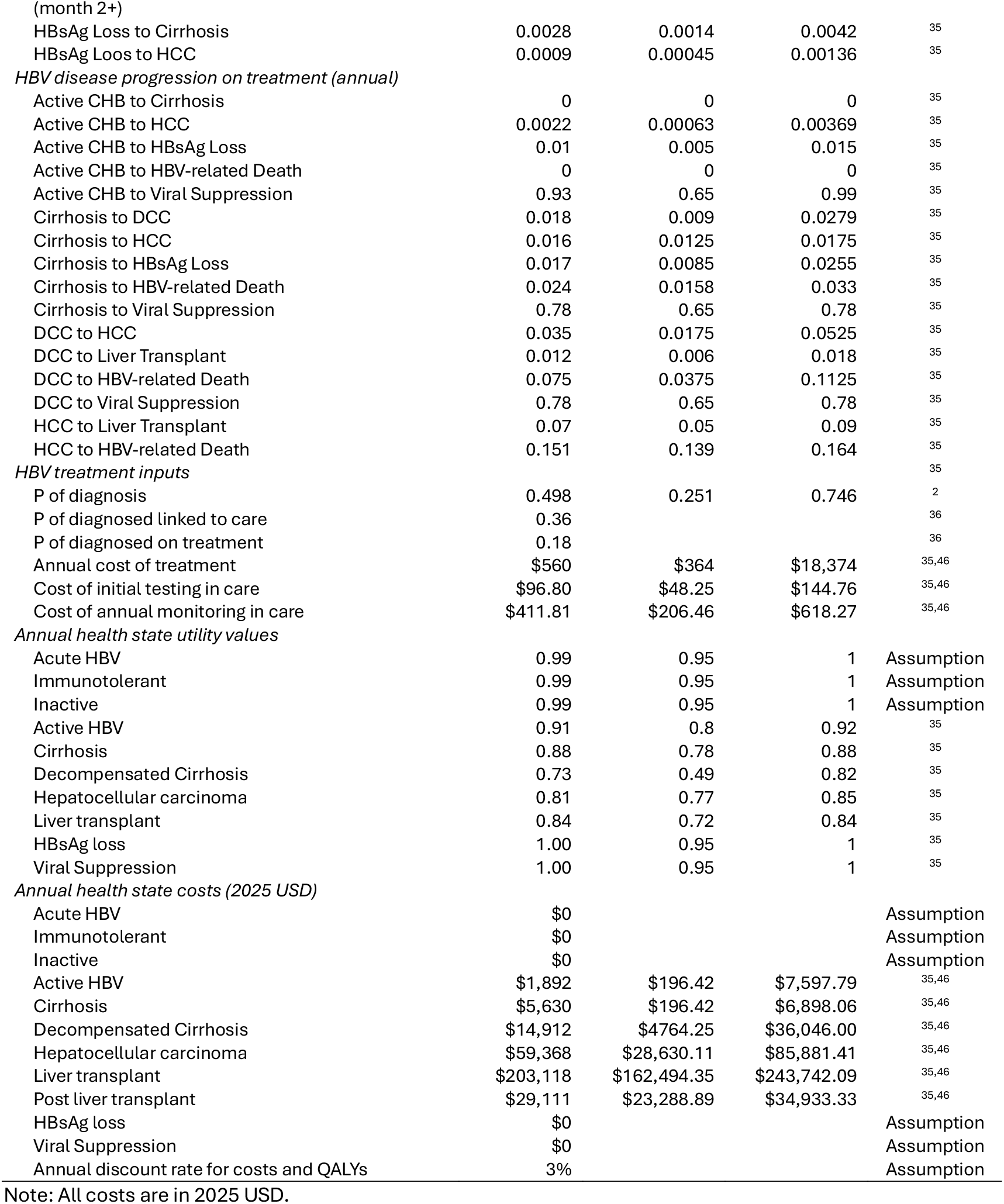
Inputs used in modeling analysis of pediatric HepB vaccination recommendations, United States, 2025.

### Vaccination

We modeled vaccination scenarios for infants with >2,000 grams birthweight and single-antigen vaccine. In all vaccination scenarios, infants born to HBsAg test-positive parents experienced a vaccination schedule consistent with current recommendations.^16^ These infants received a birth dose of HepB plus HBIG at month 0, followed by a second HepB dose at 1 month and a third HepB dose 6 months. Additionally, the comparison scenario reflected current ACIP recommendations for infants born to HBsAg test-negative and HBsAg test-unknown parents, which were operationalized in our model as a birth dose at month 0, dose 2 at month 1 and dose 3 at month 6. We modeled eight alternative vaccination scenarios that were defined by delaying the first dose by 2 months, 7 months, 4 years, or 12 years in either just the known HBsAg test-negative group or in both the HBsAg test-negative and HBsAg test-unknown group. All vaccination scenarios are described in Table 2. The HepB birth dose alone was 75% effective and the HepB birth dose plus HBIG was 94% effective in preventing perinatal transmission.^16^ For delayed vaccination series, a protective response occurred in 25% of children after dose 1, 63% after dose 2, and 98% after dose 3.^16^ We assumed vaccine efficacy did not wane during the 18 years in which modeled children were at risk of acute HBV infection.

**Table 2.** Vaccination scenarios used in modeling analysis of HepB vaccination recommendations, United States, 2025.

| Vaccination Scenarios | 1st dose | 2nd dose | 3rd dose |
| --- | --- | --- | --- |
| All scenarios |  |  |  |
| Known HBsAg+ | Birth + HBIG<br>(78.9%) | 1 month<br>(87.3%) | 6 months<br>(90.5%) |
| Comparison (current recommendations) |  |  |  |
| Known HBsAg- | Birth<br>(78.9%) | 1 month<br>(87.3%) | 6 months<br>(90.5%) |
| Unknown HBsAg status | Birth<br>(78.9%) | 1 month<br>(87.3%) | 6 months<br>(90.5%) |
| Alternative 1 - Delay 2 months among HBsAg- |  |  |  |
| Known HBsAg- | 2 months<br>(83.7%) | 3 months<br>(78.2%) | 8 months<br>(72.6%) |
| Unknown HBsAg status | Birth<br>(78.9%) | 1 month<br>(87.3%) | 6 months<br>(90.5%) |
| Alternative 2 - Delay 7 months among HBsAg- |  |  |  |
| Known HBsAg- | 7 months<br>(81.8%) | 8 months<br>(67.3%) | 13 months<br>(52.7%) |
| Unknown HBsAg status | Birth<br>(78.9%) | 1 month<br>(87.3%) | 6 months<br>(90.5%) |
| Alternative 3 - Delay 4 years among HBsAg- |  |  |  |
| Known HBsAg- | 48 months<br>(78.2%) | 49 months<br>(55.9%) | 54 months<br>(33.6%) |
| Unknown HBsAg status | Birth<br>(78.9%) | 1 month<br>(87.3%) | 6 months<br>(90.5%) |
| Alternative 4 - Delay 12 years among HBsAg- |  |  |  |
| Known HBsAg- | 144 months<br>(78.2%) | 145 months<br>(55.9%) | 150 months<br>(33.6%) |
| Unknown HBsAg status | Birth<br>(78.9%) | 1 month<br>(87.3%) | 6 months<br>(90.5%) |
| Alternative 5 - Delay 2 month among HBsAg- and unknown |  |  |  |
| Known HBsAg- | 2 months<br>(83.7%) | 3 months<br>(78.2%) | 8 months<br>(72.6%) |
| Unknown HBsAg status | 2 months<br>(83.7%) | 3 months<br>(78.2%) | 8 months<br>(72.6%) |
| Alternative 6 - Delay 7 months among HBsAg- and unknown |  |  |  |
| Known HBsAg- | 7 months<br>(81.8%) | 8 months<br>(67.3%) | 13 months<br>(52.7%) |
| Unknown HBsAg status | 7 months<br>(81.8%) | 8 months<br>(67.3%) | 13 months<br>(52.7%) |
| Alternative 7 - Delay 4 years among HBsAg- and unknown |  |  |  |
| Known HBsAg- | 48 months<br>(78.2%) | 49 months<br>(55.9%) | 54 months<br>(33.6%) |
| Unknown HBsAg status | 48 months<br>(78.2%) | 49 months<br>(55.9%) | 54 months<br>(33.6%) |
| Alternative 8 - Delay 12 years among HBsAg- and unknown |  |  |  |
| Known HBsAg- | 144 months<br>(78.2%) | 145 months<br>(55.9%) | 150 months<br>(33.6%) |
| Unknown HBsAg status | 144 months<br>(78.2%) | 145 months<br>(55.9%) | 150 months<br>(33.6%) |

Note: Values in parenthesis indicate vaccination coverage values used in scenarios that reflect imperfect recommendation adherence.

We conducted one set of analyses that assumed perfect adherence to vaccine recommendations (i.e. all persons were vaccinated according to the scenario). Additionally, we conducted a set of analyses in which only a proportion of eligible persons received each vaccine dose (referred to as imperfect adherence to recommendations). To inform these scenarios, we utilized published data from the National Immunization Survey-Child^38^ (survey of children born in 2021) and National Immunization Survey-Teen^39^ (survey year=2024), along with results from a study of HepB vaccine series completion.^40^ For adherence to scenarios that followed the current recommended timeline (i.e. birth, 1 month, and 6 months), we used published data on HepB vaccination coverage at the birth dose (79.8%), >= 2 doses (87.3%) and >=3 doses by the end of the recommended window (90.5%). For alternative vaccination schedules, we assumed the proportion of children that would start the HepB series was equivalent to published coverage estimates from other vaccination series that are recommended to be started closest to the relevant time point. For each of the modeled HepB series, we used the following initiation probabilities for each start time: 1) 2 months - Hib vaccine initiation by 3 months (83.7%), 2) 7 months - HepA vaccine initiation by 19 months (81.8%), 3) 4 years (48 months) – at least one HPV dose (78.2%), 4) 12 years (144 months) – at least one HPV dose (78.2%).^38,39^ Yusuf et al. found that earlier administration of the first HepB vaccine dose is associated with increased likelihood of completion.^40-42^ To calculate the proportion of children that complete the HepB series under each scenario, we multiplied the ratio of series completion for timing of first dose (as reported by Yusuf el al.^40^) by the proportion of children that complete the HepB series under the birth dose scenario (90.5%). Using the series initiation probabilities and series completion probabilities, we assumed a drop off between doses was equal. The proportion of children that received each dose in each imperfect adherence vaccination scenario are reported in Table 2.

### Cost and health state utilities

We conducted this analysis from the healthcare system perspective and quantified all costs related to HepB vaccination and medical management of HBV infection. We used a cost of $17.87 (range: $15.14-$29.52) for each dose of HepB vaccination, which was the published CDC price for a pediatric dose of Engerix-B as of 11/1/2025.^43^ The lower and upper bounds were the CDC cost for Recombivax and the private price for Engerix-B, respectively. For persons that received a birth dose, there were no costs associated with HBV screening prior to vaccination. Current HBV screening and testing guidelines^44^ recommend a triple panel screening test (HBsAg, anti-HBc, anti-HBs) prior to initiating HBV vaccination among adults. There is not currently an equivalent screening recommendation for vaccinating infants or children, however this would be critical to determine the HBsAg status of susceptible children prior to first vaccination. We included an additional analysis in which each person that initiated the HepB vaccination series at a time point later than the birth dose accrued the cost of a triple panel screening test (HCPCS code: G0499, $28.27 from the Centers for Medicare and Medicaid Services 2025 Q4 Clinical Laboratory Fee Schedule^45^). Medical management costs for chronic HBV infections, cirrhosis, decompensated cirrhosis, and liver cancer came from a previously published model^35^ and were adjusted for inflation to 2025 US dollars using the medical care component of the US Consumer Price Index.^46^ We assumed persons linked to care incurred costs for initial baseline tests ($96.80, range: $48.25-$144.76) and annual monitoring ($411.81 per year, range: $206.46-$618.27).^35,46^ For those that started treatment, the annual cost of antiviral drugs was $560, range: $364 - $18,374.^35,46^ Health state utilities were from the same previously published model of HBV disease progression.^35^ All costs and utilities are reported in Table 1.

## Results

All delayed vaccination scenarios resulted in an increase in HBV infections, sequelae, HBV related deaths, and total costs compared to current recommendations (Table 3). Furthermore, all delayed vaccination scenarios resulted in fewer life-years and QALYs compared to the current scenario. If assuming perfect adherence to vaccination recommendations, delaying the HepB series 2 months among children born to HBsAg test-negative parents resulted in an additional 90 acute HBV infections, 75 chronic HBV infections, 29 additional HBV-related deaths and at least $16 million ($16,428,668) in additional spending for one year of births. The number of poor health outcomes and costs increased with longer delays. Delaying the HepB series among children born to HBsAg test-negative parents to 7 months, 4 years, and 12 years resulted in an additional 90 (29 HBV-related deaths), 117 (36 HBV-related deaths) and 190 (50 HBV-related deaths) incident acute HBV infections, respectively. Furthermore, each of those scenarios resulted in an additional $16,428,668, $19,839,936, $29,916,591 in healthcare costs.

**Table 3.** Change in modeled outcomes resulting from delaying initiation of the recommended HepB vaccination schedule from birth, United States, 2025^a^.

|  | Alternative 1<br>Delay 2 months<br>HBsAg- | Alternative 2<br>Delay 7 months<br>HBsAg- | Alternative 3<br>Delay 4 years<br>HBsAg- | Alternative 4<br>Delay 12 years<br>HBsAg- | Alternative 5<br>Delay 2 months<br>HBsAg-/unknown | Alternative 6<br>Delay 7 months<br>HBsAg-/unknown | Alternative 7<br>Delay 2 years<br>HBsAg-/unknown | Alternative 8<br>Delay 12 years<br>HBsAg-/unknown |
| --- | --- | --- | --- | --- | --- | --- | --- | --- |
| Imperfect adherence to recommendations<sup>b</sup> |  |  |  |  |  |  |  |  |
| Additional acute HBV infections | 238 | 438 | 701 | 740 | 1,437 | 1,841 | 2,593 | 2,726 |
| Additional chronic HBV infections | 75 | 91 | 120 | 125 | 1,039 | 1,156 | 1,319 | 1,344 |
| Additional cirrhosis cases | 61 | 102 | 150 | 155 | 462 | 555 | 715 | 742 |
| Additional HCC cases | 39 | 68 | 103 | 107 | 304 | 370 | 484 | 503 |
| Additional HBV-related deaths | 62 | 101 | 155 | 163 | 482 | 583 | 760 | 788 |
| Additional vaccine doses | -659,734 | -1,551,463 | -2,587,254 | -2,587,254 | -837,706 | -1,974,955 | -3,293,788 | -3,293,788 |
| Number with ≥1 dose | -14,318 | -72,510 | -173,532 | -173,532 | -17,783 | -91,838 | -220,574 | -220,574 |
| Number with 3 doses | -509,325 | -1,073,737 | -1,616,540 | -1,616,540 | -647,086 | -1,366,841 | -2,058,046 | -2,058,046 |
| Change in QALYs | -12,220 | -24,255 | -39,898 | -42,373 | -61,474 | -83,812 | -129,169 | -137,342 |
| Change in life years | -1,972 | -3,174 | -5,164 | -5,512 | -17,463 | -20,831 | -27,412 | -28,303 |
| Additional costs (2025 USD) | 21,598,859 | 24,548,318 | 24,560,818 | 27,493,767 | 222,392,975 | 254,610,652 | 298,672,907 | 313,022,134 |
| Additional costs with pre-vaccination screening (2025 USD) | 123,515,177 | 124,819,549 | 121,976,157 | 124,909,106 | 324,211,338 | 354,335,480 | 394,758,368 | 409,107,595 |
| Perfect adherence to recommendations<sup>c</sup> |  |  |  |  |  |  |  |  |
| Additional acute HBV infections | 90 | 90 | 117 | 190 | 1,481 | 1,563 | 2,141 | 2,351 |
| Additional chronic HBV infections | 75 | 75 | 84 | 93 | 1,259 | 1,329 | 1,517 | 1,554 |
| Additional cirrhosis cases | 30 | 30 | 35 | 47 | 520 | 549 | 683 | 725 |
| Additional HCC cases | 21 | 21 | 26 | 37 | 328 | 350 | 451 | 481 |
| Additional HBV-related deaths | 29 | 29 | 36 | 50 | 546 | 580 | 733 | 774 |
| Additional vaccine doses | 0 | 0 | 0 | 0 | 0 | 0 | 0 | 0 |
| Number with ≥1 dose | 0 | 0 | 0 | 0 | 0 | 0 | 0 | 0 |
| Number with 3 doses | 0 | 0 | 0 | 0 | 0 | 0 | 0 | 0 |
| Change in QALYs | -3,286 | -3,286 | -4,965 | -9,265 | -58,072 | -61,205 | -95,848 | -108,471 |
| Change in life years | -1,026 | -1,026 | -1,234 | -1,783 | -19,992 | -21,149 | -26,885 | -28,265 |
| Additional costs (2025 USD) | 16,428,668 | 16,428,668 | 19,839,936 | 29,916,591 | 257,707,353 | 277,489,324 | 343,688,613 | 368,649,927 |
| Additional costs with pre-vaccination screening (2025 USD) | 119,018,633 | 119,018,633 | 122,429,901 | 132,506,555 | 360,297,318 | 380,079,288 | 446,278,577 | 471,239,891 |

<sup>a</sup> Results are based on 3,628,934 births
<sup>b</sup> All scenarios are compared to a baseline scenario of current recommendations with imperfect adherence (Table 2).
<sup>c</sup> All scenarios are compared to a baseline scenario of current recommendations with perfect adherence (Table 2).

Scenarios in which the HepB series was delayed among children born to both HBsAg test-negative and HBsAg test-unknown resulted in additional poor health outcomes and health care costs.

Assuming perfect recommendation adherence, the number of additional incident acute HBV infections was 1,481 (546 HBV-related deaths), 1,563 (580 HBV-related deaths), 2,141 (733 HBV-related deaths), and 2,351(744 HBV-related deaths) when delaying the vaccine series by 2 months, 7 months, 4 years, and 12 years, respectively. Each of those scenarios resulted in an increase in health care costs ranging from greater than $257 million (Alternative 5, delaying 2 months) to greater than $368 million (Alternative 8, delaying 12 years).

When modeling imperfect vaccination recommendation adherence, all scenarios resulted in a more pronounced increase in poor health outcomes and costs, with a reduction in life-years and QALYs. For example, delaying the HepB series among children born to HBsAg test-negative parents 2 months resulted in an additional 238 acute HBV infections, 75 chronic HBV infections, 62 HBV-related deaths and more than $21 million in additional healthcare costs. Similarly, delaying until 12 years resulted in an additional 740 acute HBV infections and greater than $24 million in health care costs. Approximately 70-90% (representing delays from 2 months to 12 years) of these excess infections are projected to occur outside the perinatal period (not shown in table). When considering imperfect recommendation adherence in scenarios that delayed HepB vaccination among both HBsAg test-negative and HBsAg test-unknown parents, the increase in health care costs ranged from over $222 million (Alternative 4, delay 2 months) to over $313 million (Alternative 8, delay 12 years).

## Discussion

Our model highlights the benefits of the current program and demonstrates added costs associated with delays in vaccine series initiation. Modeled outcomes of delaying initiation of the current ACIP infant HepB vaccination recommended immunization schedule by 2 months, 7 months, 4 years and 12 years among infants born to HBsAg test-negative parents or HBsAg test negative/unknown parents, resulted in worse outcomes (e.g., increase in HBV infections, cirrhosis, HBV related deaths), and substantially increased total costs compared to the current recommendations. When considering imperfect, more realistic, adherence to the vaccine recommendation, the negative impacts on health and costs were amplified. Furthermore, the longer the HepB series initiation was delayed in newborns and children, the higher the risk of developing chronic HBV, liver cancer, and cirrhosis, incurring more costs. For each year the HepB birth dose recommendation is delayed to 2 months for infants whose mothers are not known to be living with HBV, we anticipate at least 1,400 excess new HBV infections among children, which would incur over $222 million in additional healthcare costs. Notably, under no scenario would delaying vaccination be cost saving.

The HepB birth dose followed by a complete vaccine series provides protection against gaps in screening during pregnancy^15,16,27^, and the risk of horizontal transmission among infants and children outside of the perinatal period.^8-10^ HepB vaccines have over 40 years of robust data on efficacy and safety, demonstrating long-term immunogenicity and likely lifetime protection^47^ and post-licensure safety data has revealed no new or unexpected safety concerns over decades of review.^16,48,49^ Thus, there is no new evidence to support changing the current vaccination recommendations. To the contrary, a change risks reversing immense progress in reducing new HBV cases (99% decline among people aged 0-19 years).^19^ Whereas increasing the birth dose coverage from ∼80% to greater than 90%, rather than delaying vaccination, would likely further reduce HBV infections.^50^

Modifying the current HepB vaccine program has numerous cost implications and unintended consequences. The relatively low number of additional infections we estimate outside the perinatal period is due to the overwhelming success of the birth dose. However, there is ongoing risk of persisting low level preventable transmission (horizontal transmission). If the birth dose is delayed, the risk of community acquisition increases among future birth cohorts as population prevalence increases, resulting in more infections, adverse outcomes, and healthcare costs with each successive year. Our model indicates that approximately 70-90% (representing delays from 2 months to 12 years) excess infections that are projected to occur within childhood would occur outside the perinatal period. Therefore, our model, which projects only the impacts on one annual birth cohort, is a lower bound of the actual health and economic impacts that would be observed over time.

HBV infections are often asymptomatic for decades and historically have led to late diagnoses when advanced disease is present, thus increasing health care costs in comparison to earlier stage diagnoses.^51^ Universal adult HBV screening in the United States was implemented in 2023^44^ to promote detection of chronic HBV infections before the development of severe liver disease.

Children and adolescents aged <18 years were not included in the universal screening recommendation, though, because of the low prevalence of HBV infection in this age group and high levels of HepB vaccination, a success of the infant vaccination program. However, if initiation of the HepB vaccine series is delayed, clinical best practice would include screening children before starting the HepB vaccine series, due to their lack of vaccine protection and susceptibility to household and community exposure early in life. In our model, all scenarios increased cost from additional pre-vaccination screening among children who did not receive a birth dose.

A change to the HepB vaccination program could have profound health system implications. Though we did not model any costs specifically related to implementation, there are some implications worth considering, as they could cause substantial disruption in vaccine administration. Currently HepB birth dose vaccination is the standard of care and is operationalized as standing orders/protocols at hospitals. Experience for other infectious diseases, including for screening adults for HBV and HCV, has demonstrated that risk factor-based screening will be less effective and miss more infections that universal screening. Therefore, delaying the birth dose to after hospital discharge or later in life, could place additional burden on the primary care provider to determine vaccination history, perform risk-based screening, and potentially order diagnostic testing. If the birth dose was not recommended, places where births occur may not carry sufficient stock of vaccine, decreasing vaccine access. We reference in the model design that roughly 80% of pregnant parents whose HBsAg status was unknown at birth did not get screened at delivery.^28^ With the current guidelines, that gap in care is mitigated by the fact that all children will at least get the birth dose vaccine. Without the birth dose, it is possible that these infants will get misclassified and managed as born to HBsAg-negative parents and will face increased risk for acquiring perinatal infection. Initiating the HepB vaccine series beyond infancy results in a greater chance for missed doses in the vaccine series. In addition, receipt of the birth dose is positively associated with series completion across the childhood routine vaccinations, while delayed receipt of the HepB birth dose has negative implications for completion.^42,52,53^ A delayed birth dose recommendation may reduce vaccine confidence among HBsAg positive persons during pregnancy and has the potential to negatively influence birth dose administration in the United States and globally. Imperfect adherence (the higher cost scenario) is the more likely scenario as vaccination initiation is prolonged. In addition, if ACIP recommends a delay in vaccination where the 3 doses do not align with any of the routine childhood wellness visits^54^, then there is an even higher likelihood of missed doses and incomplete adherence.

### Limitations

Our study had several limitations. First, we assumed all infants follow vaccination schedules designed for birth weight >2,000g. Modeling scenarios where we assumed the HepB vaccine series initiation was delayed did not account for the current HepB recommendations that specify infants weighing <2,000g born HBsAg negative birth parents should get the birth dose at hospital discharge (or age 1 month if still hospitalized).^16^ As a result we likely overestimated the number of HBV infections acquired in our cohort until they receive vaccination. This overestimation, however, should be relatively small since infants born at <2,000g are likely pre-term and represent less than 10% of births in the United States.^55^ Second, we assumed all children receive single-antigen vaccine. The birth dose should always be administered as single-antigen, however subsequent doses are often administered as a combination vaccine. The cost of single-antigen vaccine is substantially less than the combination vaccines, so the cost of 1 dose might be underestimated.^43^ Third, we did not include additional costs for visits to a healthcare provider for the purpose of receiving HepB vaccine doses. Changes to the HepB vaccine schedule that require doses to be administered visits that are not part of the routine well-child visit schedule would likely incur additional costs and result in lower HepB vaccine series completion rates. Fourth, we did not model costs and health outcomes associated with serious adverse eaects from HepB vaccine.^16,48^ Except for anaphylaxis to a vaccine component, which occurs at about 1 per 1.1 million doses, there is no evidence for any other serious adverse events. Therefore, not accounting for serious vaccine adverse effects would not have substantially impacted our results. Fifth, we only modeled risk of HBV infection until 18 years of age to better isolate the costs and health outcomes associated specifically with changes to the perinatal and childhood HepB vaccine schedule and we did not assume any changes to adult HepB vaccine recommendations. As a result, our model represents a small proportion of what we would expect to see as actual population-level impact over time. The following are not represented in the model: a) incident HBV infections among the birth cohort after age 18 for those who remain unvaccinated, b) onward transmissions from infections acquired among the modeled birth cohort or future birth cohorts, the c) increasing risk of horizontal transmission (community and household acquisition) among future birth cohorts as population prevalence increases, resulting in more infections, adverse outcomes, and healthcare costs with each successive year. Importantly, our approach was conservative, and the limitations based on our model assumptions underestimate costs and health outcomes associated with delays to administering the HepB birth dose. All limitations appreciably affecting model outputs would result in even greater impact of delayed HepB birth dose if addressed.

## Conclusion

Our model demonstrates that maintaining the current ACIP HepB vaccine recommendations (i.e., administration of the birth dose <24 hours for all infants, regardless of birthing parent HBsAg-status, followed by completion of the HepB vaccine series) prevents the most number of perinatal and childhood HBV infections and is cost saving to the health system compared with any scenario that results in delays to initiating the HepB vaccine series. Aligning with the World Health Organization’s 2009 recommendation to incorporate the birth dose into national childhood vaccination programs ensures that the United States is adhering to global standards for best practice and will ensure progress towards meeting national and global HBV elimination goals.

Delaying the HepB birth dose in the United States would result in worse outcomes, increased costs, and a reversal of progress in the United States towards elimination^56^ of chronic HBV infection, an incurable disease.

## Data Availability

All data produced in the present study are available upon reasonable request to the authors

## References

1. Wong RJ, Brosgart CL, Welch S, et al. An Updated Assessment of Chronic Hepatitis B Prevalence Among Foreign-Born Persons Living in the United States. Hepatology (Baltimore, Md). Mar 3 2021; doi:10.1002/hep.31782

2. Bixler D, Barker L, Lewis K, Peretz L, Teshale E. Prevalence and awareness of Hepatitis B virus infection in the United States: January 2017 - March 2020. Hepatol Commun. Apr 1 2023;7(4) doi:10.1097/hc9.0000000000000118

3. Kowdley KV, Wang CC, Welch S, Roberts H, Brosgart CL. Prevalence of chronic hepatitis B among foreign-born persons living in the United States by country of origin. Hepatology (Baltimore, Md). Aug 2012;56(2):422–33. doi:10.1002/hep.24804

4. Abara WE, Qaseem A, Schillie S, McMahon BJ, Harris AM. Hepatitis B Vaccination, Screening, and Linkage to Care: Best Practice Advice From the American College of Physicians and the Centers for Disease Control and Prevention. Annals of internal medicine. Dec 5 2017;167(11):794–804. doi:10.7326/m17-1106

5. Beasley RP, Hwang LY, Lin CC, Chien CS. Hepatocellular carcinoma and hepatitis B virus. A prospective study of 22 707 men in Taiwan. Lancet (London, England). Nov 21 1981;2(8256):1129–33. doi:10.1016/s0140-6736(81)90585-7

6. Goldstein ST, Zhou F, Hadler SC, Bell BP, Mast EE, Margolis HS. A mathematical model to estimate global hepatitis B disease burden and vaccination impact. International journal of epidemiology. Dec 2005;34(6):1329–39. doi:10.1093/ije/dyi206

7. McMahon BJ, Alberts SR, Wainwright RB, Bulkow L, Lanier AP. Hepatitis B-related sequelae. Prospective study in 1400 hepatitis B surface antigen-positive Alaska native carriers. Archives of internal medicine. May 1990;150(5):1051–4. doi:10.1001/archinte.150.5.1051

8. Sabeena S, Ravishankar N. Horizontal Modes of Transmission of Hepatitis B Virus (HBV): A Systematic Review and Meta-Analysis. Iran J Public Health. Oct 2022;51(10):2181–2193. doi:10.18502/ijph.v51i10.10977

9. West DJ, Margolis HS. Prevention of hepatitis B virus infection in the United States: a pediatric perspective. The Pediatric infectious disease journal. Oct 1992;11(10):866–74. doi:10.1097/00006454-199210000-00012

10. Davis LG, Weber DJ, Lemon SM. Horizontal transmission of hepatitis B virus. Lancet (London, England). Apr 22 1989;1(8643):889–93. doi:10.1016/s0140-6736(89)92876-6

11. Bond WW, Favero MS, Petersen NJ, Gravelle CR, Ebert JW, Maynard JE. Survival of hepatitis B virus after drying and storage for one week. Lancet (London, England). Mar 7 1981;1(8219):550–1. doi:10.1016/s0140-6736(81)92877-4

12. McMahon BJ, Bulkow LR, Singleton RJ, et al. Elimination of hepatocellular carcinoma and acute hepatitis B in children 25 years after a hepatitis B newborn and catch-up immunization program. Hepatology (Baltimore, Md). Sep 2 2011;54(3):801–7. doi:10.1002/hep.24442

13. Hyams KC. Risks of Chronicity Following Acute Hepatitis B Virus Infection: A Review. Clinical Infectious Diseases. 1995;20(4):992–1000. doi:10.1093/clinids/20.4.992

14. Centers for Disease Control and Prevention (CDC). Prevention of perinatal transmission of hepatitis B virus: prenatal screening of all pregnant women for hepatitis B surface antigen. MMWR Morbidity and mortality weekly report. Jun 10 1988;37(22):341–6, 351.

15. Willis BC, Wortley P, Wang SA, Jacques-Carroll L, Zhang F. Gaps in hospital policies and practices to prevent perinatal transmission of hepatitis B virus. Pediatrics. Apr 2010;125(4):704–11. doi:10.1542/peds.2009-1831

16. Schillie S, Vellozzi C, Reingold A, et al. Prevention of Hepatitis B Virus Infection in the United States: Recommendations of the Advisory Committee on Immunization Practices. MMWR Recommendations and reports : Morbidity and mortality weekly report Recommendations and reports. 2018;67(RR-1):1–31. doi:10.15585/mmwr.rr6701a1

17. Centers for Disease Control and Prevention (CDC). Update: recommendations to prevent hepatitis B virus transmission--United States. MMWR Morbidity and mortality weekly report. Aug 4 1995;44(30):574–5.

18. Centers for Disease Control and Prevention (CDC). Update: recommendations to prevent hepatitis B virus transmission--United States. MMWR Morbidity and mortality weekly report. Jan 22 1999;48(2):33–4.

19. Bixler D, Roberts H, Panagiotakopoulos L, Nelson NP, Spradling PR, Teshale EH. Progress and Unfinished Business: Hepatitis B in the United States, 1980-2019. Public health reports (Washington, DC : 1974). Jun 9 2023:333549231175548. doi:10.1177/00333549231175548

20. Centers for Disease Control and Prevention (CDC). National Notifiable Diseasea Surveillance System (NNDSS). Accessed November 17, 2023. https://www.cdc.gov/nndss/index.html

21. World Health Organization. Global health sector strategy on viral hepatitis 2016-2021. Towards ending viral hepatitis. 2016.

22. World Health Organization. Hepatitis B vaccines. Weekly epidemiological record. 2 October 2009 2009;No. 40(84):405–420.

23. Martin JA, Hamilton BE, Osterman MJK. Births in the United States, 2024. NCHS data brief. Jul 2025;(535):1. doi:10.15620/cdc/174613

24. Kushner T, Chen Z, Tressler S, Kaufman H, Feinberg J, Terrault NA. Trends in Hepatitis B Infection and Immunity Among Women of Childbearing Age in the United States. Clinical infectious diseases : an oXicial publication of the Infectious Diseases Society of America. Jul 27 2020;71(3):586–592. doi:10.1093/cid/ciz841

25. Schillie S, Walker T, Veselsky S, et al. Outcomes of infants born to women infected with hepatitis B. Pediatrics. May 2015;135(5):e1141–7. doi:10.1542/peds.2014-3213

26. Martin JA, Osterman MJK. Changes in Prenatal Care Utilization:United States, 2019-2021. National vital statistics reports : from the Centers for Disease Control and Prevention, National Center for Health Statistics, National Vital Statistics System. May 2023;72(4):1–14.

27. Pham TTH, Maria N, Cheng V, et al. Gaps in Prenatal Hepatitis B Screening and Management of HBsAg Positive Pregnant Persons in the U.S., 2015-2020. American journal of preventive medicine. Jul 2023;65(1):52–59. doi:10.1016/j.amepre.2023.01.041

28. Euler GL, Wooten KG, Baughman AL, Williams WW. Hepatitis B surface antigen prevalence among pregnant women in urban areas: implications for testing, reporting, and preventing perinatal transmission. Pediatrics. May 2003;111(5 Pt 2):1192–7.

29. Abbott Laboratories. ARCHITECT HBsAg Next Qualitative Reagent IFU https://www.accessdata.fda.gov/cdrh_docs/pdf21/P210003C.pdf

30. Roche Diagnostics. Elecsys HBsAg II Package Insert. https://www.fda.gov/media/176636/download

31. McMahon BJ, Alward WL, Hall DB, et al. Acute hepatitis B virus infection: relation of age to the clinical expression of disease and subsequent development of the carrier state. The Journal of infectious diseases. Apr 1985;151(4):599–603.

32. Balogun MA, Parry JV, Mutton K, et al. Hepatitis B virus transmission in pre-adolescent schoolchildren in four multi-ethnic areas of England. Epidemiology and infection. May 2013;141(5):916–25. doi:10.1017/s0950268812001513

33. Nelson NP, Jamieson DJ, Murphy TV. Prevention of Perinatal Hepatitis B Virus Transmission. J Pediatric Infect Dis Soc. Sep 2014;3 Suppl 1(Suppl 1):S7–s12. doi:10.1093/jpids/piu064

34. Fan L, Owusu-Edusei K, Jr., Schillie SF, Murphy TV. Cost-effectiveness of active-passive prophylaxis and antiviral prophylaxis during pregnancy to prevent perinatal hepatitis B virus infection. Hepatology (Baltimore, Md). May 2016;63(5):1471–80. doi:10.1002/hep.28310

35. Toy M, Hutton D, Harris AM, Nelson N, Salomon JA, So S. Cost-Effectiveness of 1-Time Universal Screening for Chronic Hepatitis B Infection in Adults in the United States. Clinical infectious diseases : an oXicial publication of the Infectious Diseases Society of America. Jan 29 2022;74(2):210–217. doi:10.1093/cid/ciab405

36. Harris AM, Osinubi A, Nelson NP, Thompson WW. The hepatitis B care cascade using administrative claims data, 2016. The American journal of managed care. Aug 2020;26(8):331–338. doi:10.37765/ajmc.2020.44069

37. Arias E, Xu J, Kochanek K. United States Life Tables, 2022. National vital statistics reports : from the Centers for Disease Control and Prevention, National Center for Health Statistics, National Vital Statistics System. Apr 8 2025;(2)

38. Centers for Disease Control and Prevention (CDC). ChildVaxView. Accessed November 8, 2025. https://www.cdc.gov/childvaxview/index.html

39. Centers for Disease Control and Prevention (CDC). TeenVaxView. Accessed November 8, 2025. https://www.cdc.gov/teenvaxview/index.html

40. Yusuf HR, Daniels D, Smith P, Coronado V, Rodewald L. Association between administration of hepatitis B vaccine at birth and completion of the hepatitis B and 4:3:1:3 vaccine series. Jama. Aug 23-30 2000;284(8):978–83. doi:10.1001/jama.284.8.978

41. Hawkins SS. Long-term Implications and Barriers to Use of the Hepatitis B Vaccine at Birth. J Obstet Gynecol Neonatal Nurs. Nov 2024;53(6):594–606. doi:10.1016/j.jogn.2024.09.008

42. Oster NV, Williams EC, Unger JM, et al. Hepatitis B Birth Dose: First Shot at Timely Early Childhood Vaccination. American journal of preventive medicine. Oct 2019;57(4):e117–e124. doi:10.1016/j.amepre.2019.05.005

43. Centers for Disease Control and Prevention (CDC). Current CDC Vaccine Price List. https://www.cdc.gov/vaccines-for-children/php/awardees/current-cdc-vaccine-price-list.html

44. Conners EE, Panagiotakopoulos L, Hofmeister MG, et al. Screening and Testing for Hepatitis B Virus Infection: CDC Recommendations - United States, 2023. MMWR Recommendations and reports : Morbidity and mortality weekly report Recommendations and reports. Mar 10 2023;72(1):1–25. doi:10.15585/mmwr.rr7201a1

45. Center for Medicare & Medicaid Services (CMS). 25CLABQ4. https://www.cms.gov/medicare/payment/fee-schedules/clinical-laboratory-fee-schedule-clfs/files/25clabq4

46. Bureau of Labor Statistics (BLS). Consumer Price Index. https://www.bls.gov/cpi/

47. Bruce MG, Bruden D, Hurlburt D, et al. Protection and antibody levels 35 years after primary series with hepatitis B vaccine and response to a booster dose. Hepatology (Baltimore, Md). Oct 2022;76(4):1180–1189. doi:10.1002/hep.32474

48. Haber P, Moro PL, Ng C, et al. Safety of currently licensed hepatitis B surface antigen vaccines in the United States, Vaccine Adverse Event Reporting System (VAERS), 2005-2015. Vaccine. Jan 25 2018;36(4):559–564. doi:10.1016/j.vaccine.2017.11.079

49. Eriksen EM, Perlman JA, Miller A, et al. Lack of association between hepatitis B birth immunization and neonatal death: a population-based study from the vaccine safety datalink project. The Pediatric infectious disease journal. Jul 2004;23(7):656–62. doi:10.1097/01.inf.0000130953.08946.d0

50. Hill HA, Yankey D, Elam-Evans LD, Chen M, Singleton JA. Vaccination Coverage by Age 24 Months Among Children Born in 2019 and 2020 - National Immunization Survey-Child, United States, 2020-2022. MMWR Morbidity and mortality weekly report. Nov 3 2023;72(44):1190–1196. doi:10.15585/mmwr.mm7244a3

51. Le MH, Liu JK, Lee K, Cheung R, Nguyen MH. Late Diagnosis of Chronic Hepatitis B in the United States: A Population-Based, Retrospective Cohort Study From 2007 to 2021. Alimentary pharmacology & therapeutics. Aug 2025;62(3):340–348. doi:10.1111/apt.70193

52. Vader DT, Lee BK, Evans AA. Hepatitis B Birth Dose Effects on Childhood Immunization in the U.S. American journal of preventive medicine. Feb 2020;58(2):208–215. doi:10.1016/j.amepre.2019.10.007

53. Wilson P, Taylor G, Knowles J, et al. Missed hepatitis B birth dose vaccine is a risk factor for incomplete vaccination at 18 and 24 months. J Infect. Feb 2019;78(2):134–139. doi:10.1016/j.jinf.2018.09.014

54. American Academy of Pediatrics. AAP Schedule of Well-Child Care Visits. Accessed November 2023, 2025. https://www.healthychildren.org/English/family-life/health-management/Pages/Well-Child-Care-A-Check-Up-for-Success.aspx

55. Martin J, Osterman MJK. Trends in Early-term Singleton Births in the United States, 2014 to 2023. NCHS Health E Stats. National Center for Health Statistics (US) All material appearing in this report is in the public domain and may be reproduced or copied without permission; citation as to source, however, is appreciated.; 2024.

56. National Academies of Sciences E, Medicine. A National Strategy for the Elimination of Hepatitis B and C: Phase Two Report. In: Strom BL, Buckley GJ, eds. A National Strategy for the Elimination of Hepatitis B and C: Phase Two Report. National Academies Press (US) Copyright 2017 by the National Academy of Sciences. All rights reserved.; 2017.

